# Zapping the brain to enhance sport performance? Evidence from an umbrella review of the effect of transcranial direct current stimulation on physical performance

**DOI:** 10.1101/2024.03.07.24303915

**Authors:** Darías Holgado, Daniel Sanabria, Miguel A. Vadillo, Rafael Román-Caballero

## Abstract

Concepts such as "neuro-doping" or brain doping have contributed to an expansion in the area of transcranial direct current stimulation (tDCS) and its impact over exercise and physical performance in recent years. Here we assess the evidence supporting the healthy population using an umbrella review of meta-analyses investigating the role of tDCS to enhance exercise performance. We identified 9 meta-analyses encompassing 50 crossover studies and 683 participants that met our inclusion criteria. Despite the fact that most meta-analyses reported a positive effect of tDCS, our analyses revealed overly low statistical power in the primary studies, publication bias, and large variability in pre-processing and analytic decisions. Indeed, a specification-curve analysis showed that the final effect could range from *g* = −0.23 to *g* = 0.33, depending on decisions such as the formula used for estimating the effect size and multiple additional analytic steps. Moreover, a meta-analysis of all the primary studies included in the umbrella review showed a small effect of tDCS (*g*_z_ = 0.28, 95%CI [0.18, 0.39]) that became substantially smaller and inconclusive after accounting for publication bias, *g*_rm_ = 0.10, 95%CrI [−0.04, 0.20], *BF*_10_ = 0.99. In summary, our findings highlight that current evidence, from both individual studies and meta-analyses, does not conclusively support the idea that tDCS enhances performance outcomes.

## Introduction

In the pursuit of optimizing performance, exercise scientists explore avenues for marginal gains, a trend exacerbated by the escalating pressure on athletes to continually enhance their abilities. A recently prominent method garnering attention from athletes across various sports is transcranial direct current stimulation (tDCS; (1). The underlying premise behind tDCS usage is its potential to elevate physical performance by stimulating specific brain regions (e.g., motor or prefrontal cortex) engaged during exercise. While the precise mechanisms through which tDCS may enhance physical exercise performance remain elusive, the current hypothesis attributes potential improvements to factors such as diminished pain perception, heightened corticospinal excitability, and a reduction in perceived effort during exercise (2).

The increasing interest in this technique in recent years is evident from the escalating number of primary studies (3–5), narrative reviews (6), systematic reviews (7), and meta-analyses (8–10) published in the last few years. Although the focus of these studies and meta-analyses might vary slightly, the main conclusions of the literature are that (1) tDCS has an ergogenic effect, albeit a small one; (2) tDCS might be more effective in some exercise performance domains than others; (3) the level of expertise of the population might be relevant (i.e., novice participants might have more room for improvement); (4) little is known of the long-term effect of the stimulation; (5) the effects may be modulated by various factors (e.g., the area stimulated, the electrode placement, the duration of stimulation, etc.).

This umbrella review delves into the current landscape of tDCS and its impact on exercise performance. The overarching objective is to discern the validity of claims asserting tDCS as a beneficial ergogenic aid, evaluating whether these assertions are substantiated by robust evidence or merely represent a transient trend in the field.

## Methods

### Pre-registration

The methods and planned analyses of this umbrella review were pre-registered on 29 December 2022 at PROSPERO (CRD42022384967) and in the OSF repository: https://osf.io/73qsu/. All major deviations from the pre-registered protocol and analysis plans are transparently identified in the manuscript.

### Data and code availability

The data analyzed and the code used for the analysis in this study are publicly available at the OSF repository: https://osf.io/73qsu/.

#### Literature search

We conducted a systematic literature search following the Preferred Reporting Items for Systematic Reviews and Meta-Analyses guidelines (last search in January 2023) in Medline, Web of Science and Scopus using the following Boolean operators: ("physical exercise" OR "exercise performance” OR "physical activity" OR sport) AND (tDCS OR tES OR "brain stimulation" OR “transcranial stimulation”) AND (meta-analysis OR metaanalysis OR “systematic review”). Additionally, we searched on Google Scholar to identify unpublished meta-analyses meeting the inclusion criteria. Search was limited to papers published in English.

#### Inclusion and exclusion criteria

We followed the Participant-Intervention-Comparison-Outcome process to select the meta-analyses included in this umbrella review: (1) Participants: healthy participants of all ages and both sexes. (2) Intervention: within and between studies investigating the effects of tDCS on physical exercise performance. (3) Comparison: anodal or cathodal stimulation vs. sham control. (4) Outcome: meta-analyses and primary studies should report at least one measure of physical exercise performance. Upon review, we observed that most of the literature is composed of primary studies with within-participant manipulations. We advance here that only 3 of the 53 studies that met the inclusion criteria involved between-group designs (11–13). For the sake of simplicity, we decided a posteriori to restrict the analyses to the 50 studies with within-participant designs, although the conclusions were not affected when the three between-group studies were included (**Supplementary Material 1**).

### Data Extraction

The following data were extracted from each meta-analysis by DH and RRC: (1) list of authors and year of publication from each primary article included in the meta-analysis; (2) latest search date and publication date; (3) type and estimation method of effect size; (4) reported final effect size; (5) method for dealing with dependence between effect sizes; (6) type of exercise outcome and stimulation analyzed; (7) number of included studies; (8) analysis of publication bias; and (9) protocol registration. In addition, for our assessment of transparency and reproducibility practices, we coded whether the meta-analysis reported compliance with reporting guidelines, competing interests, search limits, search terms, full search strategy, eligibility criteria, double coding, use of methods to assess risk of bias in primary studies, dealing with dependence between effect sizes and the combination of between- and within-participant designs, outlier identification, statistical model, estimation method of the heterogeneity variance, software, and code and data availability.

At the primary study level, DH, DS, MAV and RRC extracted the following information from the studies included in the meta-analyses in duplicated: (1) list of authors and year of publication; (2) pooled number of participants for the stimulation and sham group; (3) sample characteristics; (4) type of exercise test; (5) exercise category; (6) exercise outcome assessed; (7) study design; (8) target brain area, electrode montage, stimulation duration and intensity, electrode surface, number of sessions, and concurrence of the stimulation; (10) *t* or *F* value; and (11) means and standard deviations for each condition (when we could not extract exact means and standard deviations but graphic information was available, we estimated the values from the graphs using the software WebPlotDigitizer; (14). Based on these data, we identified 50 primary studies that met the criteria of the present umbrella review. Moreover, the effect sizes were only extracted for the comparisons of interest. Therefore, effect sizes comparing conditions with and without supplementation (e.g., caffeine) or assessing the impact of other factors than tDCS (e.g., mental fatigue) were excluded.

#### Statistical analysis

##### Graph analysis

To explore the overlap in primary studies across the reviewed meta-analyses, we created a bipartite network graph (with meta-analyses and primary studies as two different categories of nodes) using the *igraph* R package (15). To analyze the centrality and closeness of the nodes, we converted the two-mode network into two one-mode networks and, for meta-analyses, we used the *tnet* R package (16) to calculate the sum of weights on ties originating from a node as *centrality score* and the normalized *closeness score* for nodes (dividing raw closeness score by *N* − 1).

##### Effect size

Given that most of the primary studies adopted a study design in which the physical performance of the same sample of participants was compared after tDCS vs. a sham condition (within-participant designs contrasting posttest performance), we opted for a Cohen’s *d*_z_, which we preferentially estimated from *t* and *F* values 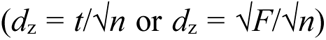. If those values were not available (note that with *F* value we mean the statistic for the main effect of stimulation in a repeated measures ANOVA with only two tDCS conditions), we used the following formula as a standardized mean difference-based effect size:

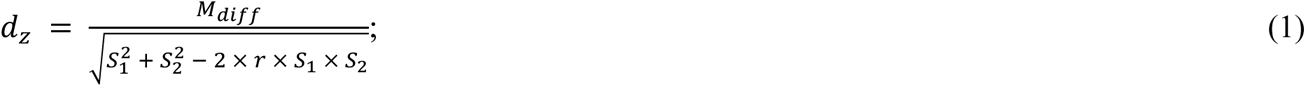

where *M*_diff_ represents the difference of means, *S*_1_ and *S*_2_ the standard deviations of the contrasted conditions, and *r* the repeated-measures correlation. Its variance was estimated as:

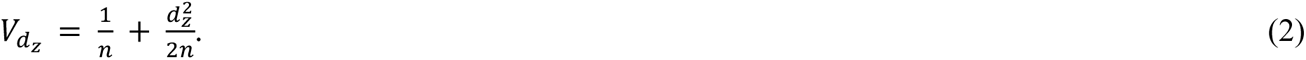

Note that this decision was taken considering that the intra-individual difference score is the most natural unit of analysis in this type of research, which aims to prove the causal effect of tDCS using the participant themself as a control for other background variables (for a more detailed discussion, see (17). Therefore, we excluded a minority of three studies with between-group designs (see **Inclusion and exclusion criteria**) and estimated *d*_z_ in the remaining studies. We estimated repeated-measures correlations from *t* values and *F* values from one-way repeated measures ANOVA (i.e., 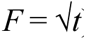) to reach an overall repeated-measures correlation that could be imputed in studies that did not report any of both statistics. Overall, we could extract 44 correlations from 26 studies (mean of 1.7 correlations per study, 1–6) and, subsequently, we obtained a meta-analytic Pearson’s *r* of .846, 95%CI [.76, .93]. For studies in which means and standard deviations could not be extracted, *d*_z_ was calculated directly from the *t* value and the number of participants: 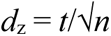. Additionally, we provide a transformation of that effect into *common language effect* (*CL*; (18); i.e., the probability that a randomly sampled score from the treatment condition is greater than another score sampled from the control condition).

Although the main analyses reported in our umbrella review were based on studies with within-participant designs, all the analyses were repeated including the three studies that implemented interventions between groups (**Supplementary Material 1**). To combine effects across within and between-participant designs, we used *d*_rm_ and *d*_s_ formulas for within- and between-participant designs, respectively:

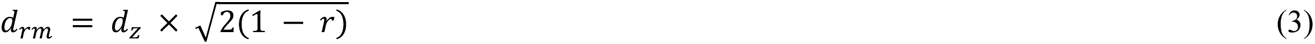

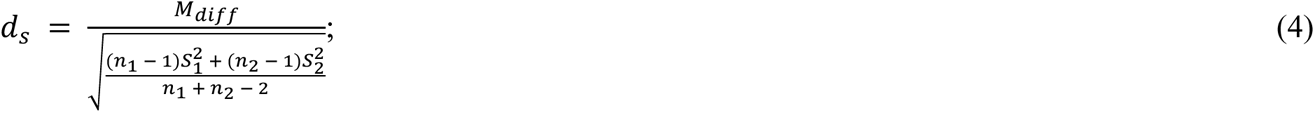

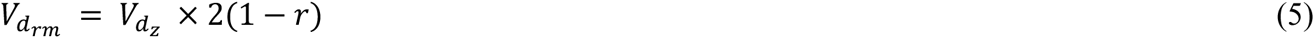

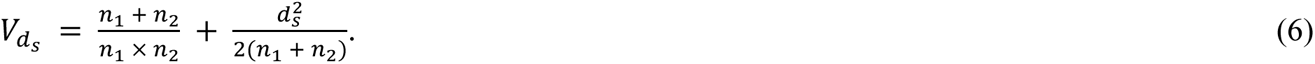

All estimates and their variance were corrected for small-sample bias:

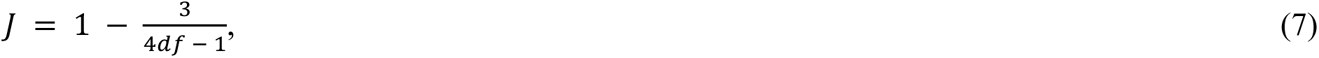

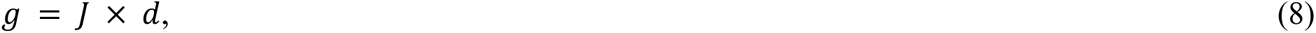

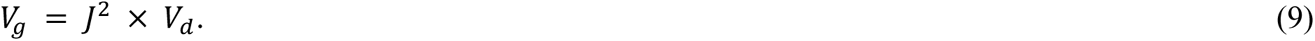

##### Meta-analysis and heterogeneity

We implemented multilevel meta-analytic models using the robust variance estimation approach (RVE; (19), which deals with a correlated structure of outcomes from the same primary study. We used the RVE method using the *robumeta* R package (20). The usual heterogeneity indexes, τ^2^ and *I*^2^, were computed.

##### Outlier detection and moderator analysis

We assessed whether the observed heterogeneity could be due to the presence of outliers and moderating variables. We fitted a multilevel model with the *rma.mv* function of *metafor* (21) and estimated the studentized residuals. Studies with studentized residuals higher than 2 were identified as outliers. In addition, as already planned in the registration of this study, we examined the influence of the following moderators: (1) target brain area (motor, prefrontal, or temporal cortex); (2) stimulation duration (in min; continuous variable); (3) stimulation intensity (in mA; continuous variable); (4) concurrence of the stimulation with the exercise task (online, offline or offline-online); (5) type of outcome (endurance or strength exercise); and (6) muscle involved (whole body vs. isolated)^1^. Moreover, we investigated the role of further moderators (previously not included in the registered protocol): (1) year of publication (continuous variable); (2) age of the sample (in years; continuous variable); (3) training status (untrained vs. trained); (4) stimulation polarity (cathodal vs. anodal); (5) electrode montage (single vs. bicephalic vs. high definition, HD); (6) electrode surface (in mm; continuous variable); (7) return location (extracephalic vs. cephalic); and (8) number of sessions (continuous variable).

##### Statistical power and publication bias

To estimate the power of individual studies, we took as reference an approximation of our final uncorrected effect (*g*_z_ = 0.30), and found out the number of participants needed to reach specific power thresholds, assuming a one-tailed *t*-test and an alpha of .05. To test for publication bias, we relied on two types of methods, based either on funnel plot asymmetry (FAT) or selection models. Among the first category is the inclusion of the effect-size precision estimate as a moderator in the multilevel model to test whether there is a general relationship between the observed effect sizes and their precision (i.e., funnel plot asymmetry; (22). Within this procedure, the intercept of the meta-regressive model would be taken as the best estimate of the underlying effect (i.e., the estimated effect when the sampling error is zero), a method that has been proposed to follow a conditional procedure (*precision-effect test– precision-effect estimate with standard error*, PET–PEESE; (23). The logic of PET-PEESE can be extended to multilevel models (24–26). We conducted FAT and PET-PEESE with Fisher’s *z* for being a variance-stabilizing transformation for the effect size and preventing the artifactual dependence between Cohen’s *d* and its precision estimate (27). For the Fisher’s *z* transformation, we converted *d*_z_ into *d*_rm_ (17), and then *d*_rm_ into Fisher’s *z* (28).

Within the second type, selection models (29) assume that the probability of publication depends on the *p* value. In our meta-analysis, we used a three-parameter selection model (3PSM) with a one-tailed *p*-value cutpoint of .025, selecting only significant studies. A way to account for dependence among effect sizes with this model is to combine all the effect sizes coming from the same sample generating an average estimate for each study, and conduct the classic methods on these aggregates (26). We used the *MAd* package in R (30) to generate within-study aggregates, while we carried out 3PSM with the *weightr* package (31).

Finally, we conducted a robust Bayesian meta-analysis (RoBMA; (32) that yields one single model-averaged estimate after simultaneously applying (1) meta-regressive models for the relationship between effect sizes and their standard errors (PET–PEESE) and (2) selection models that estimate relative publication probabilities (selection model). RoBMA makes inferences guided mostly by those models that predict the observed data best. Based on the reviewed meta-analyses that reflect a prior belief of a substantial effect of tDCS on exercise performance, we selected a normal distribution centered at 0.30 and with one standard deviation as the prior of the effect in the alternative hypothesis. For the effect belonging to the null hypothesis, we assumed a normal distribution centered at 0 an equal standard deviation. As with the FAT and PET-PEESE method and 3PSM, RoBMA was conducted with a Fisher’s transformation of *d*_z_ and within-study aggregates.

To reduce the impact of heterogeneity on the output of publication-bias analyses, we drew out known heterogeneity due to outliers and moderators (stimulation intensity and polarity) in all the methods. Thus, we excluded outlying studies and outcomes coming from cathodal tDCS (which represented a minority) and added stimulation intensity as a continuous moderator (re-centered at 0). In the case of RoBMA, we first fitted a univariate meta-analysis with the aggregates and stimulation intensity as a moderator, from which the raw residuals plus the intercept of that model served as the input for RoBMA.

##### Specification curve

Finally, we conducted an exploratory specification curve analysis (33) with all the primary studies. In the specification curve, we estimated the final effect size and its significance for a total of 32 possible combinations of four analytic decision levels: (a) how to deal with within-study dependence (no strategy or assuming within-effects independence, an RVE multilevel model, and fitting a univariate model with aggregate effect sizes); (b) the identification and exclusion or not of outlying studies; (c) the inclusion or not of influential moderators to adjust the outcome (that is, type of control group and baseline difference); and (d) the strategies to assess and correct the final outcome for publication bias (PET-PEESE, 3PSM, and no correction). Although all meta-analyses estimated the effect size following the *g*_s_ formula, we conducted the specification-curve analysis adopting three formulas of the effect size for comparative purposes: *g*_rm_, *g*_s_, and *g*_z_. The analysis led to 96 different combinations of specifications, as 3PSM cannot be conducted with a multilevel model. Six models did not converge properly (all of them with 3PSM, and *g*_s_ or *g*_z_), which led to a total of 90 outcomes.

## Results

A total of 9 meta-analyses (8–10, 34–39) meeting the inclusion criteria were selected from among the 1,145 records retrieved in the search. We identified 70 primary studies in the meta-analyses, of which

50 met the inclusion criteria for our umbrella review (see **Supplementary Material 2** for exclusion reasons), involving a total of 683 participants. We extracted 101 effect sizes from them, a mean of two outcomes per study (range 1–6). Most of the primary studies used offline tDCS (45), only anodal stimulation (42), and young samples of participants (48; between 16.1 and 33 years old). The studies applied tDCS over the primary motor cortex (36), prefrontal cortex (11), temporal cortex (6), or the cerebellum (1) to investigate its benefits on strength and endurance exercise performance.

### Overlapping and variation in study sampling

The graphical visualization of the connections between the meta-analyses and primary studies (**Figure 1**) showed a large overlap in a significant portion of the meta-analyses (6 out of 9). The average centrality score (53.2, 95%CI [40.5, 65.8]) and the normalized closeness scores (1.14, 95%CI [0.96, 1.33]) of this group differ from the indices of the other three remaining meta-analyses (10, 35, 37; centrality score: 11, 24, and 10; normalized closeness score: 0.40, 0.62, and 0.39), as evidence of this distancing from the central group (**Table 1**). The separation of these meta-analyses could be due to divergences in the inclusion criteria they used, especially regarding the selection of participants. While most of the meta-analyses only required that participants should be healthy adults, the works by Alves-Lobão et al. (35) and Maudrich et al. (10) constrained their reviews to studies with samples of athletes. Hu et al. (37), on the contrary, selected tDCS interventions with untrained adults. Alves-Lobão et al. (35) additionally restricted their search to articles from 2009 onwards. Finally, these meta-analyses are among the most recent and, therefore, included recent articles that earlier reviews missed due to a temporal reason. Other divergences in the age criteria of participants (some meta-analyses included older adults; (34, 36), stimulation polarity (some meta-analyses included cathodal stimulation; (9, 34–36), and the inclusion of only endurance and sprint performance during cycling and running tasks in Kaushalya et al. (39) had less impact on the overlap of meta-analyses^2^. At the primary study level, although the reviewed meta-analyses included an average of 15 primary studies (4–34), a large proportion of primary studies only appeared in one meta-analysis (19 out of 50). This denotes that the central group of reviews was built from sharing few primary studies (only 20 primary studies appeared at least in three reviews).

**Figure 1.**
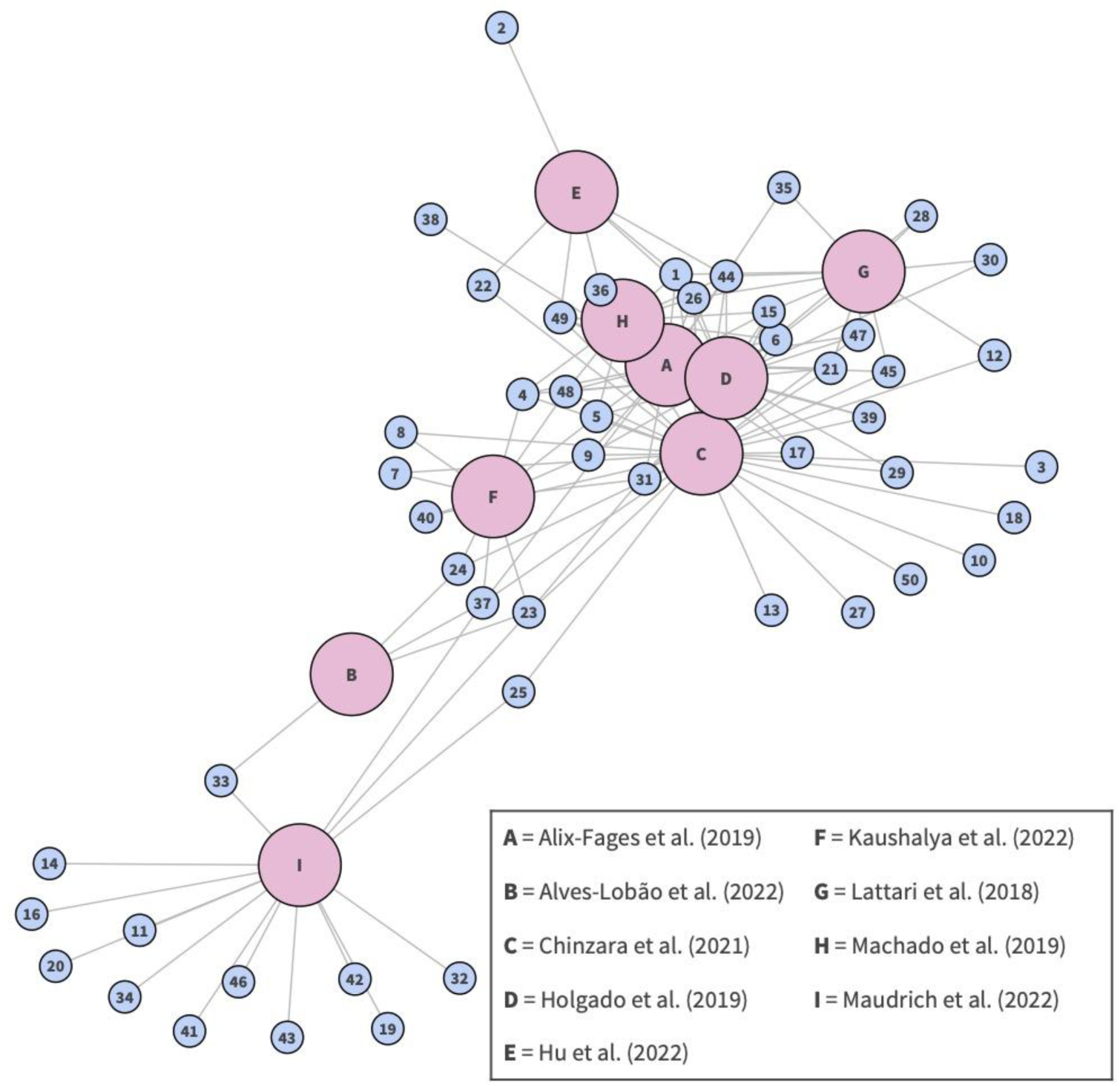
Network graph of the reviewed meta-analyses. Meta-analysis nodes are represented by letters (A–I), while the included primary studies are depicted by numerical nodes (1–49).

**Table 1.** Results of network analyses.

| Meta-analysis | Centrality score | Closeness score |
| --- | --- | --- |
| Alix-Fages et al. (2019) | 66 | 1.35 |
| Alves-Lobão et al. (2022) | 11 | 0.40 |
| Chinzara et al. (2021) | 78 | 1.50 |
| Holgado et al. (2019) | 66 | 1.35 |
| Hu et al. (2022) | 24 | 0.62 |
| Kaushalya et al. (2022) | 31 | 0.85 |
| Lattari et al. (2018) | 35 | 0.87 |
| Machado et al. (2019) | 43 | 0.96 |
| Maudrich et al. (2022) | 10 | 0.39 |

### Transparency and reproducibility practices

Before analyzing the results of meta-analyses quantitatively, we explored their transparency and reproducibility practices (40). Most of the meta-analyses included statements of compliance with reporting guidelines (8 out of 9; **Figure 2**), were free of competing interests (8), indicated the search limits (8), the search terms used (9), the full search strategy (exact terms and the Boolean connectors; 9), the eligibility criteria (9), described in details the collection process of study characteristics (9), methods to assess risk of bias in included studies (9), stated the statistical model assumed for the synthesis process (9), and identified the software used to carry out the analyses (9)^3^. However, few meta-analyses pre-registered their protocols (2; Alves-Lobão et al., 2022; Machado et al., 2019) and, although some of them used a double coding strategy (5), none reported a measure of inter-coder agreement. Two meta-analyses based their quantitative synthesis on unstandardized mean differences (Hu et al., 2022; Machado et al., 2019), combining measures from a different scale, while the remaining meta-analyses used Cohen’s *d* as the estimate of the effect size. Despite the disparity of formulas proposed for calculating Cohen’s *d* depending on the error term used for standardization (17), only two meta-analyses specified the formula in the article (39) or in the scripts of analyses (8). Most of the meta-analyses did not report how they dealt with correlated structures of effects (i.e., multiple outcomes from the same sample; 2) and how they combined effect sizes from between- and within-participant designs (1 out of 7). They also failed to describe sensitivity analyses to assess the effect of outliers (2), state the estimation method of the heterogeneity variance (1), assess the impact of publication bias (4); and make available their scripts of analyses when R packages were used (1 out of 3). Finally, even when most of the meta-analyses reported some raw data in the paper (8), in most of the cases this report was in the article itself and not in a machine-readable format (1).

**Figure 2.**
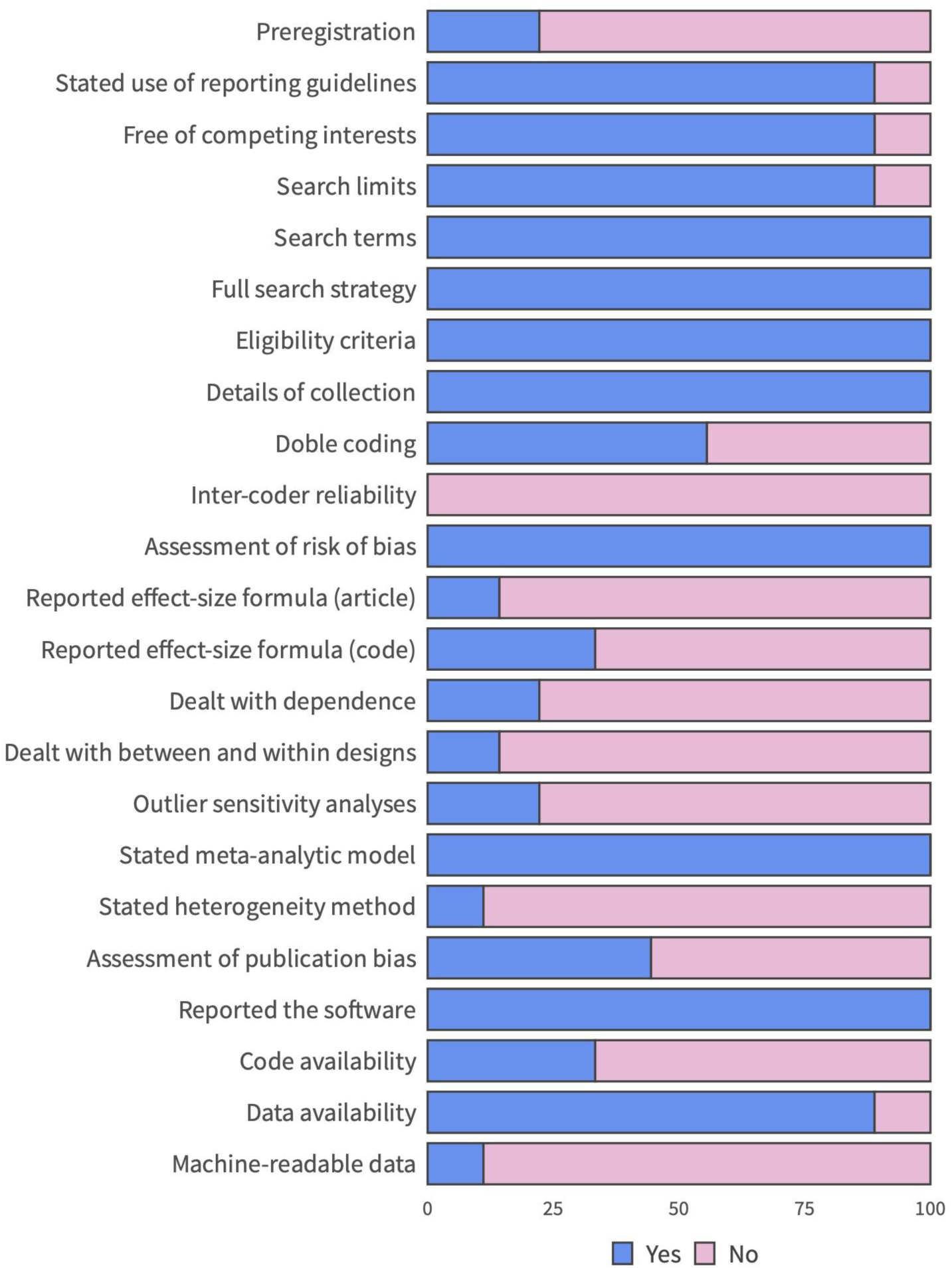
Percentage of meta-analyses according to their transparency and reproducibility practices.

### Overall effect of tDCS on exercise performance

The seven meta-analyses that evaluated the effect of tDCS using standardized effect sizes (i.e., (9, 37) used unstandardized mean differences) showed on average an effect of 0.44 (0.22 – 1.44). However, among these meta-analyses, the extremely disparate value of the review by Alves-Lobão and collaborators (35); 2022; *g* = 1.44) makes this work an outlier. Excluding this meta-analysis, the average meta-analytic effect was reduced substantially: *g* = 0.27 (0.22–0.34). This outcome is similar in magnitude to the overall effect obtained in our own meta-analysis using a multilevel model with all primary studies included from the nine meta-analyses. We observed an overall effect of *g*_z_ = 0.28, 95%CI [0.18, 0.39], *p* < .0001, *CL* = .58, and moderate heterogeneity, *I*^2^ = 55.63% (**Figure 3A**).

**Figure 3.**
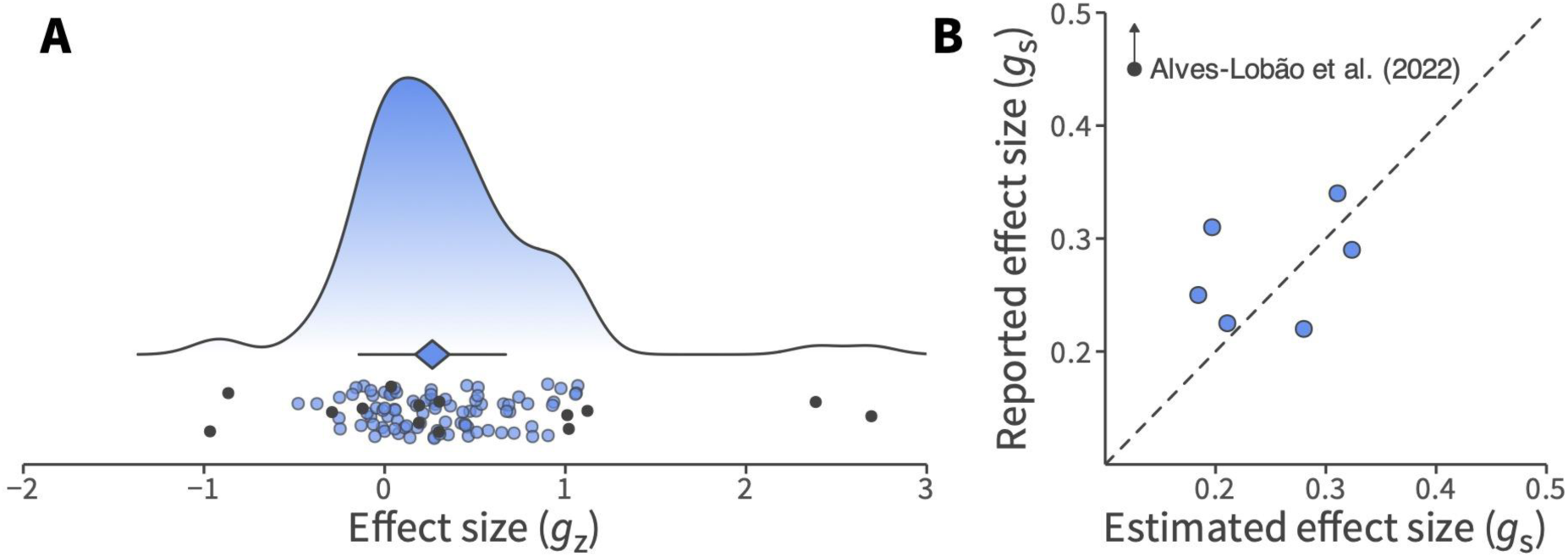
(**A**) Overall effect of the model with the individual effects of all the included primary studies. The final effect and its confidence interval are represented by the location and the width of the diamond, while the horizontal line depicts the prediction interval. Filled circles are the individual effects, while the black ones represent outlier studies. (**B**) Relationship between the reported effect sizes of the reviewed meta-analyses and the effect sizes estimated in our reanalysis. The dashed diagonal indicates the perfect match between the two estimates.

Since multiple decisions could produce variability between the reviewed meta-analyses and ours (e.g., the way the individual effect sizes were estimated or the strategy adopted to deal with the dependence generated by the inclusion of several outcomes from the same sample), we re-estimated the overall effect of each meta-analysis with the same original samples of primary studies, excluding primary studies that did not meet our inclusion criteria this time. On average, the seven reported standardized effects departed 0.23 *g*_s_ units from their corresponding re-estimated effects (0.05 after excluding Alves-Lobão and collaborators, (35); **Figure 3B**).

### Outliers and moderating variables

The moderate heterogeneity observed in our multilevel meta-analysis with all primary studies (*I*^2^ = 56.46%) might be due to the presence of outliers or other sources of variability, such as moderating variables. Five outlying studies were detected (4, 41–44) as they contributed with extremely large effect sizes (i.e., *g*_z_ < −0.8 or > 2; see dark points in **Figure 2a**). After excluding those studies, heterogeneity reduced substantially (*I*^2^ = 34.08%) while the overall effect size remained significant, *g*_z_ = 0.26, 95%CI [0.17, 0.36], *p* < .0001, *CL* = .57. In addition, two factors explained part of the between-studies variability: stimulation polarity and intensity (**Table 2**). While anodal tDCS showed a positive effect on exercise performance, *g*_z_ = 0.28, 95%CI [0.19, 0.38], *p* < .0001, cathodal tDCS did not have a significant impact, *g*_z_ = 0.03, 95%CI [−0.19, 0.25], *p* = .713 (**Figure 4A**). This result was supported by a numerical trend toward a greater improvement in performance with anodal stimulation compared to cathodal stimulation in studies where the two tDCS protocols were applied to the same sample: *g*_z, anodal vs. cathodal_ = 0.39, 95%CI [−0.08, 0.86], *p* = .081. For stimulation intensity, higher mA produced a higher effect, *p* = .021 (**Figure 4B**).

**Figure 4.**
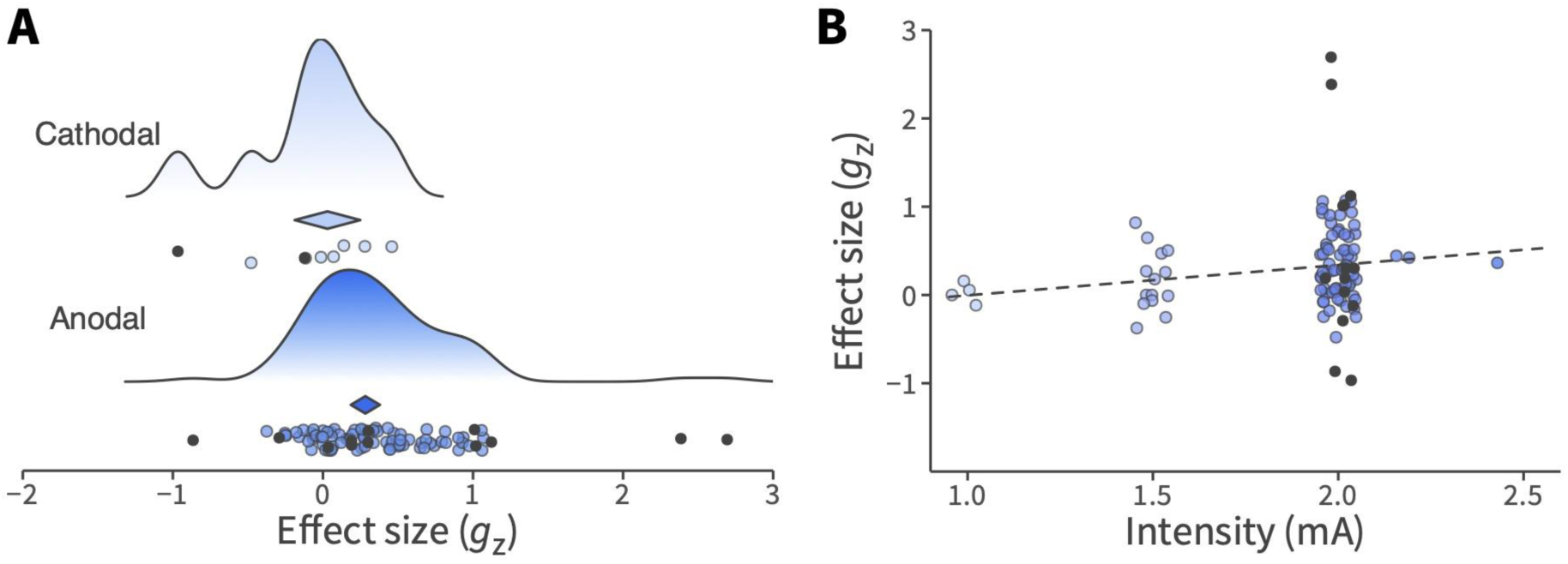
(**A**) Difference between the effects of anodal and cathodal tDCS on exercise performance. The final effect and its confidence interval are represented by the location and the width of the diamond, respectively. (**B**) Increase in the effect of tDCS with higher stimulation intensity. The dashed line denotes the trend estimated by the model. In both panels, filled circles are the individual effects, while the black ones represent outlier studies.

**Table 2.**
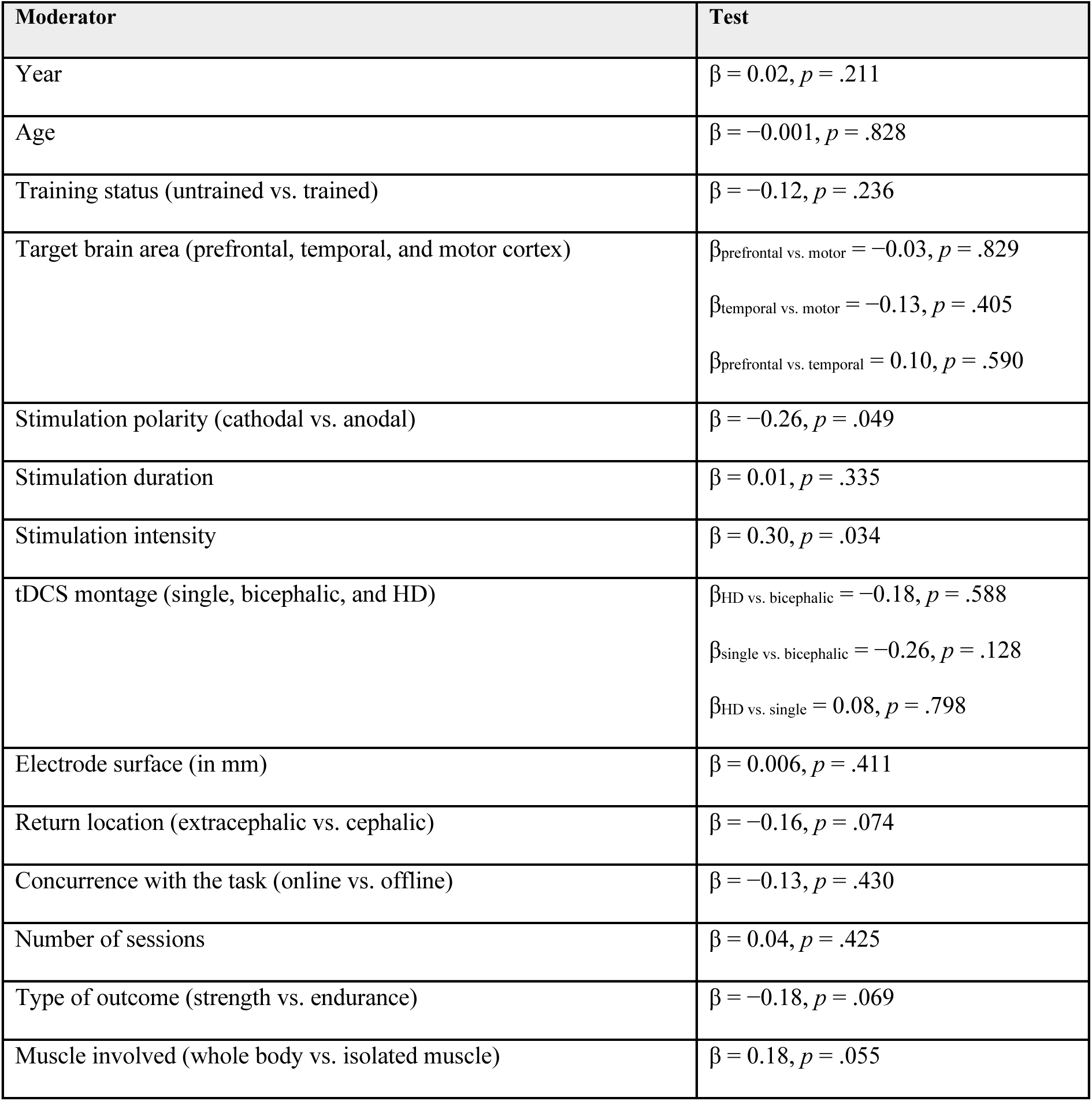
Results of moderator analyses.

### Power analysis and publication bias

The mean number of participants per study was 13 participants, a sample size that would largely stand with insufficient statistical power (< 80%) to test in a one-tailed contrast for a target effect such as the observed effect in our comprehensive meta-analysis (approximated to *g*_z_ = 0.30 for the sake of simplicity). For an effect of that size, most of the studies achieved a power of less than 40% (48 out of 50; **Figure 5A**) and only one study reached a power larger than 50%, (3); 54%). One of the reasons for this limitation could have been the grounding of the primary studies’ power analysis on the outcomes of other studies with disproportionately large effects (e.g., (11, 45, 46). Thus, in a closer examination of the accumulation of evidence in this literature, it can be observed that the average final effect of the published meta-analyses and the effects of individual studies have become increasingly closer to the effect observed in our meta-analysis over the years (**Figure 5B**). However, this evidence has not been translated to proper power analyses in the primary studies. The studies that performed power analyses to estimate their sample sizes (20 out of 50) have selected increasingly larger target outcomes over time and, whereas meta-analyses and accumulated evidence suggested an uncorrected effect size of 0.30, primary studies have based their estimation on larger effect sizes (approximated to *g*_z_ = 0.80, on average). In addition to reducing the likelihood of observing a significant effect if the effect truly exists, low statistical power also reduces the likelihood that a statistically significant result reflects a true effect (47). When a small study observed a significant and large effect, it is more probable that the result did not represent a true effect and that its estimated magnitude was inflated.

**Figure 5.**
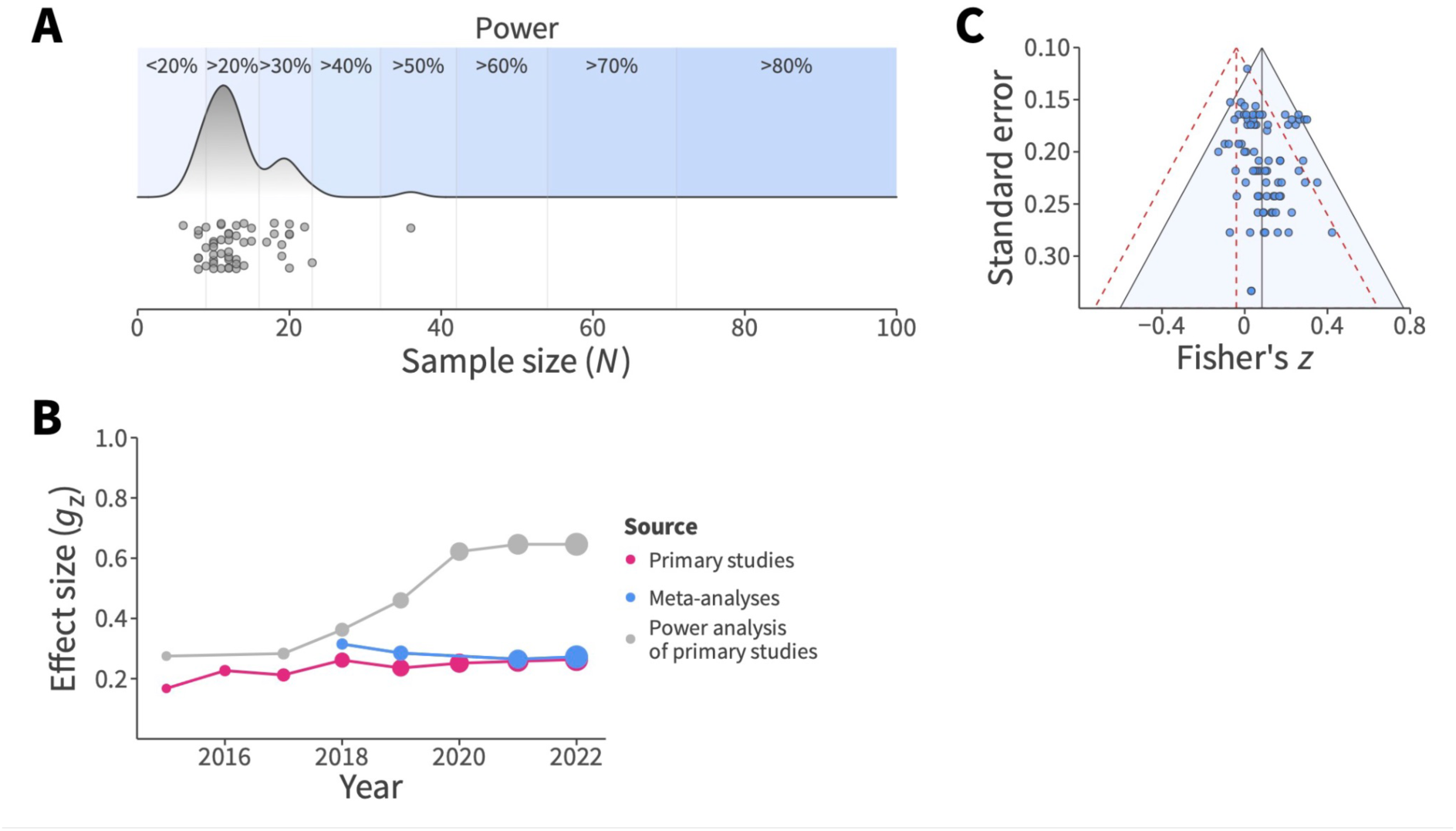
(**A**) Sample size distribution of the included primary studies and the achieved statistical power for an effect of *g*_z_ = 0.30 and a one-tailed test. (**B**) Evolution of the average final effect from primary studies (estimated by our multilevel meta-analyses over the years), the average final effect of the published meta-analyses, and the reference effects used by primary studies to conduct power analyses. (**C**) Funnel plot of the individual effects. The altitude of the solid funnel denotes the unadjusted effect, while the altitude of the dashed funnel denotes the adjusted one. Filled circles represent the individual effects.

Complementarily, we assessed publication bias after taking into account the identified sources of heterogeneity (i.e., outliers and moderators). We tested for FAT with a multilevel model that included standard error as an additional moderator. Our FAT model showed a numerical trend of standard error to explain part of the remaining heterogeneity (β = 0.62, *p* = .056; **Table 3**) and the adjusted effect of tDCS on exercise performance became non-significant, Fisher’s *z* = −0.04, 95%CI [−0.18, 0.10], *p* = .539, *CL* = .48 (**Figure 5C**). In contrast, 3PSM did not detect significant evidence of publication bias, χ^2^(1) = 1.00, *p* = .317, and the publication-bias corrected effect was significant, *g*_z_ = 0.21, 95%CI [0.04, 0.37], *p* = .017, *CL* = .56. However, when we applied robust Bayesian meta-analysis to integrate both publication bias assessment approaches into a single model-averaged estimate, the model suggested moderate evidence of publication bias, *BF*_pb_ = 3.10, and inconclusive evidence of a positive effect, *BF*_10_ = 0.99, for a posterior mean estimate of Fisher’s *z* = 0.05, 95%CrI [−0.02, 0.10], *CL* = .53.

**Table 3.** Results of methods of publication bias.

| Method | Publication bias test | Corrected effect |
| --- | --- | --- |
| FAT and PET-PEESE | $\beta = 0.64, p = .056$ | Fisher's $z = -0.04$, 95%CI $[-0.18, 0.10]$, $p = .539$ (or $g_z = -0.08$, 95%CI $[-0.37, 0.20]$, $CL = .48$) |
| 3PSM | $\chi^2(1) = 1.00, p = .317$ | $g_z = 0.21$, 95%CI $[0.04, 0.37]$, $p = .017$, $CL = .56$ |
| RoBMA | $BF_{pb} = 3.10$ | Fisher's $z = 0.05$, 95%CrI $[-0.02, 0.10]$, $BF_{10} = 0.99$ (or $g_{rm} = 0.10$, 95%CrI $[-0.04, 0.20]$, $CL = .53$) |

### Influence of analytical decisions

The differences in the multiple analytic steps the meta-analyses adopted could largely influence their outcomes. The meta-analyses differed, for example, in the approach used to deal with the within-effects dependence, whether they addressed (or not) outlying studies, the use (or not) of influential moderators and methods for assessing publication bias to adjust the final effect. To examine the impact of all these decisions on the meta-analytic outcome, we conducted an exploratory (not pre-registered) specification curve analysis. The analysis revealed that the final effect could vary greatly depending on the effect size formula used and analytic decisions (from performance impairment, *g* = −0.23, to performance enhancement, *g* = 0.33; **Figure 6**). Common specifications in the reviewed meta-analyses, such as not dealing with within-study dependence using univariate models (8 out of 9), not identifying outliers (7), or not correcting for publication bias (5) led to higher effects and more likely to be significant. In general, the uncorrected effect of tDCS was larger when the formula used was *g*_z_ instead of *g*_rm_ or *g*_s_ because *g*_z_ takes into account the correlation between repeated measures (i.e., higher between-measure correlation leads to a decrease in the standard deviation of the difference score used to standardize and a subsequent increase of *d* for the same difference score). Therefore, our specification curve analysis highlighted the large impact of analytic decisions on the final effect and that most of the meta-analyses opted for specifications that tend to find more positive outcomes.

**Figure 6.**
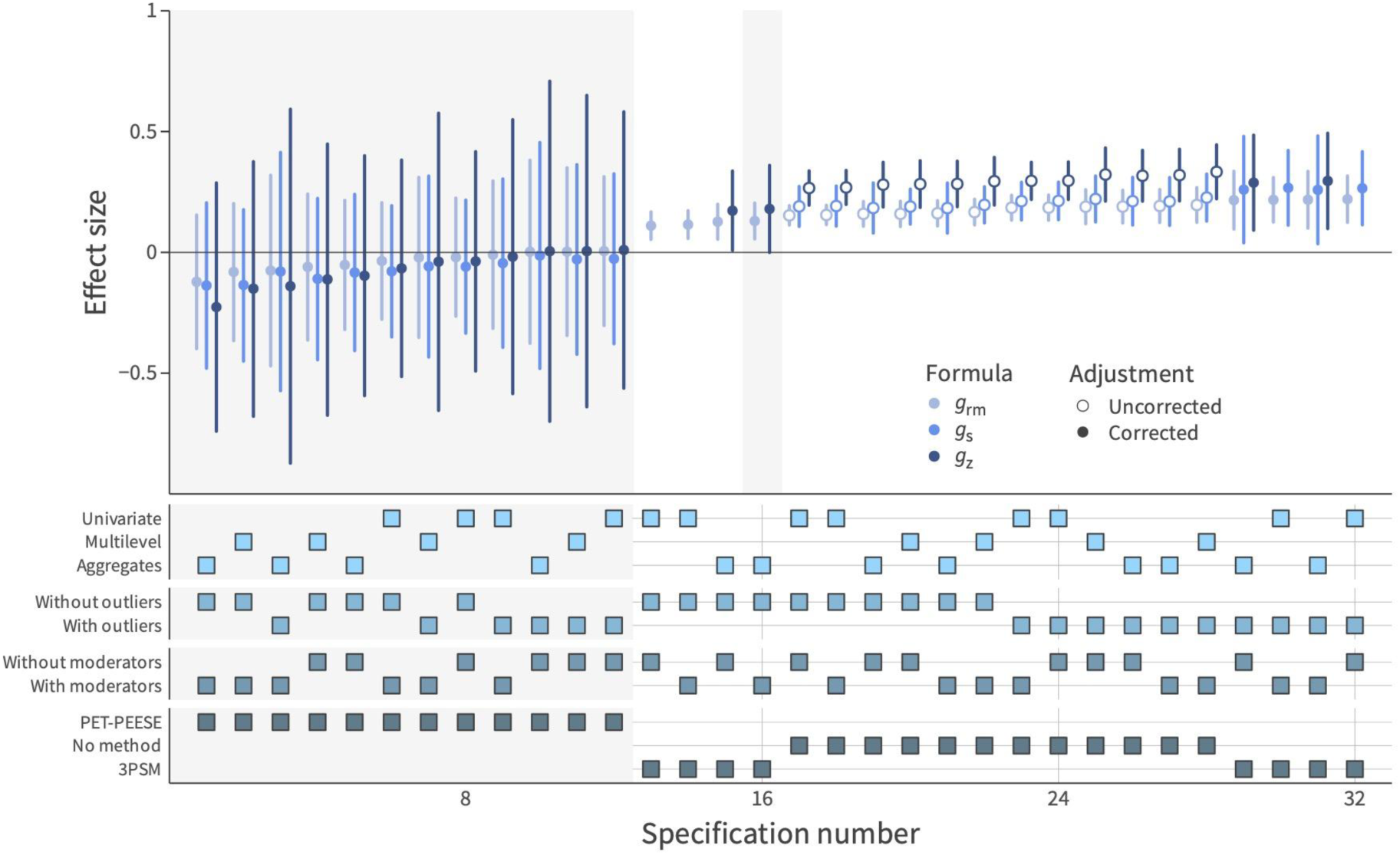
Specification curve of meta-analytic models. The summary effect size of the target studies (and its 95%CI) varied across the multiple combinations of analytic decisions. Empty circles represent the effects resulting from models without publication-bias adjustment, whereas filled circles show the corrected summary effect. Light-gray shades distinguish non-significant results from significant ones.

## Discussion

Within this umbrella review, we scrutinized the assertion that tDCS holds a potential ergogenic impact on exercise performance. Through a comprehensive re-analysis of 9 meta-analyses, comprising 50 individual studies, encompassing 101 effect sizes, and involving 683 participants, there was no conclusive evidence to support the hypothesis that transcranial brain stimulation results in discernible physical performance benefits in healthy adults. Our findings underscore the notion that the accelerated proliferation of studies and meta-analyses in this domain in recent years lacks the corresponding rigor and substantive evidence needed to definitively support the purported ergogenic effects on exercise performance. We have identified that the inconclusive evidence stems from both theoretical and methodological factors, a clarification of which is provided below.

From a theoretical point of view, the rapid advancement of this brain stimulation technology has brought the concept of “brain-doping” to the forefront of discussions (1, 48, 49). Nevertheless, the mechanisms underlying the purported effects remain unclear, with much of the impact ascribed merely to tDCS influencing the brain. A critical issue with this assertion lies in its foundational assumption that specific brain areas play a fundamental role in exercise performance. Although it might seem intuitive, the existence of a specific brain region responsible for regulating effort, task execution, or the perception of effort is not conclusively established yet, and the evidence remains unclear (50–52). The majority of primary studies in this field have utilized motor cortex stimulation to enhance performance with the foundational premise that this form of stimulation holds the potential to heighten corticospinal excitability, thereby amplifying the neural drive to muscles (2). This enhancement, in turn, is anticipated to optimize the ability for force generation while concurrently postponing the onset of fatigue or reducing pain perception (41). In contrast, other multiple studies chose the prefrontal cortex as the stimulation site (3, 53). Certain models propose that the prefrontal cortex serves as a regulatory framework that consolidates information encountered during physical activity, both centrally and peripherally, exercising top-down control (54). The prefrontal cortex is suggested to integrate afferent signals from the anterior cingulate cortex and the orbitofrontal cortex, associated with motivational and emotional processing (55). Nevertheless, the potential implications for performance remain uncertain when considering whether merely augmenting excitability or inhibiting pain in a typical state can have a discernible impact (56). Another aspect that has so far eluded extensive discussion in the literature, yet could bear notable significance, pertains to the participant’s condition. Conventionally, stimulation is administered during periods of rest, before exercise when fatigue is absent or there is no sensation of pain. An intriguing possibility arises—could stimulation exert a more substantial effect post-exercise? This consideration speculates on the prospect of stimulation serving as an excitability mechanism for accelerated recovery, potentially enabling individuals to perform more promptly following physical exertion (or during exhaustive exercise).

Given the inconclusive nature and divergent outcomes associated with tDCS, there has been a proliferation of alternative approaches proposed to enhance its efficacy. Many unanswered questions persist, including considerations about the optimal timing of stimulation, the participant’s cognitive state, and the most effective intensity. Criticism within this technique arises, as doubts are cast on the effectiveness of single-electrode stimulation. The uncertainty lies in how the electrical current disperses across the entire skull (57). To overcome this constraint, the utilization of HD-tDCS has been suggested as a means to enhance focus during stimulation. This approach aligns with the principles of traditional tDCS but employs additional electrodes with reduced sizes and precise placements to intricately target specific brain regions. However, exploration of this avenue in the literature remains limited, with only four studies included in our umbrella review employing this montage and our moderator analysis did not show that the montage impacted the overall effect. Likewise, insights from related fields suggest that conducting stimulation concurrently with task performance, known as online stimulation, might enhance its effects, benefiting on the brain’s increased susceptibility during task engagement (58, 59). In other words, when the brain is actually engaged in the target task is when one might expect an effect (60). Nevertheless, within the scope of this comprehensive analysis, only five primary studies have explored this particular aspect and, our moderator analysis did not indicate any discernible influence from this factor.

From a methodological perspective, we first delve into our exploratory analysis of the transparency and reproducibility practices of the meta-analyses included in this umbrella review and their potential impact on the results (40). Although the decision concerning how the effect size is estimated might have an enormous impact on the results and conclusions of meta-analyses, only 2 of 9 meta-analyses included the formula used to calculate the effect size (8, 39). This issue was also underscored in the amalgamation of effect sizes without accounting for study design distinctions, such as between-versus within-participant designs. Moreover, many meta-analyses incorporated multiple outcomes from the same sample, yet they did not specify how correlated structures from the same participants were addressed or how diverse outcomes were synthesized. Our specification curve analysis emphasizes that meta-analyses involve methodological choices that can significantly impact the ultimate results. By exploring various model specifications, we can gather insights into the robustness of our findings. The chosen decision can lead to a wide-ranging summary effect, ranging from indicating a negative impact of tDCS on exercise performance to suggesting a positive effect. However, most of the previous meta-analyses chose specifications for data preprocessing and analyses that are more prone to find a positive effect (i.e., lack of assessment of publication bias, no consideration of the influence of outliers and moderator studies to account for heterogeneity and conduct analyses of publication bias after removing these sources of variability, failure to deal with dependence between effect sizes from the same sample, and the use of a between-groups formula of the effect size for within-participant designs). It is evident that the conclusions drawn from these meta-analyses are constrained by these methodological considerations.

Furthermore, it is important to note that while meta-analyses might offer a valuable approach to addressing the issue of low statistical power, they should not be taken as the ultimate answer to the debate. The effectiveness of meta-analyses in overcoming the limitations of individual studies heavily relies on the quality of the reports incorporated into the analysis. Indeed, a well-designed, adequately powered, and thoroughly researched individual study holds the potential to provide more insightful and reliable information to determine the causal role of tDCS under specific conditions than the collective body of meta-analyses published so far. However, our power analysis has unveiled a concerning trend within the individual studies encompassed by our umbrella review. On average, these studies demonstrated less than 40% statistical power to detect the purported summary effect size (approximated to *g*_z_ = 0.30). This issue can be attributed to various factors. Notably, a mere 20 out of the 50 studies included in our review conducted a power analysis (or any other justification, refer to **Supplementary Material 3**). Moreover, many of them selected extreme effect sizes (e.g., (11, 45, 46) as a reference for their power analysis, even when for some cases reviews and meta-analyses suggesting a substantially smaller effect (e.g., (38) were available by that moment. Another noteworthy issue we identified is the misapplication of effect types. Some primary studies utilized metrics like η^2^ or Cohen’s *d* to determine sample size, despite their experimental designs not aligning well with such measures. This mismatch suggests a potential source of inconsistency in the reported power and, consequently, the reliability of the findings.

In summary, our findings underscore that the current body of evidence from both individual studies and meta-analyses exploring the impact of tDCS on exercise performance does not conclusively substantiate the notion that tDCS holds the potential to enhance exercise performance outcomes. In the most optimistic scenario, the estimated effect stands at *g_z_* = 0.28 (*CL* = .58; i.e., the probability that a randomly sampled score from the tDCS condition is greater than the sham condition is 8% above the chance level), a value that undergoes substantial reduction upon adjusting for publication bias. However, the inherent low power to detect this effect across all primary studies included in this review raises skepticism about the practical utility of tDCS. While the field has witnessed a surge of interest in recent years, it remains challenging to unequivocally affirm its usefulness. To address this uncertainty, it is imperative to develop more refined hypotheses and methodologies for future investigations in this domain.

### Author contributions

D.H., M.A.V., R.R.-C., and D.S. were involved in the original conceptualization. D.H., M.A.V., R.R.-C., and D.S. were responsible for developing the study methodology. D.H. did the literature search. D.H., M.A.V., R.R.-C., and D.S were responsible for data curation. R.R.-C. did the formal statistical analysis. R.R.-C. and D.H. wrote the original draft. M.A.V. and D.S. edited and reviewed the manuscript.

### Competing interests

The authors declare no competing interests.

Correspondence and requests for materials should be addressed to Darías Holgado or Rafael Román

## Data Availability

Data is available in https://osf.io/73qsu/

## Acknowledgements

This research was supported by a research grant from the Spanish Ministry of Science and Innovation to D.S. (PID2019-105635GB-I00) and from the State Research Agency to M.A.V. (CNS2022-135346). The funders had no role in study design, data collection and analysis, decision to publish or preparation of the manuscript

## Footnotes

1 Our combined analysis with within- or between-participant designs (using *g*_rm_ and *g*_s_ as estimates of the effect size, respectively) showed no difference between the effects of both study designs, β_within vs. between_ = −0.10, *p* = .714.

2 The fact that only two studies were conducted with older adults and that all cathodal tDCS interventions were applied in studies that also used anodal tDCS meant that the divergences of criterias in age and type of intervention did not result in differences in study samples.

3 Note that most of the reviewed meta-analyses (6) used Review Manager (Cochrane Collaboration, Oxford, UK; (9, 10, 34, 37–39), while three reviews relied on R packages (8, 35, 36). We restricted the assessment of code availability and the report of the effect-size formula in the code to these three latter meta-analyses.

