## Supplementary Material 1 for "Zapping the brain to enhance sport performance? Evidence from an umbrella review of the effect of transcranial direct current stimulation on physical performance"

**Reasons of Primary Study Exclusion**

**A. No exercise performance outcome**

Tanaka, S., Hanakawa, T., Honda, M., & Watanabe, K. (2009). Enhancement of pinch force in the lower leg by anodal transcranial direct current stimulation. *Experimental Brain Research, 196*, 459–465.

Seidel, O., & Ragert, P. (2019). Effects of transcranial direct current stimulation of primary motor cortex on reaction time and tapping performance: A comparison between athletes and non-athletes. *Frontiers in Human Neuroscience, 13*, 103.

Seidel-Marzi, O., & Ragert, P. (2020). Anodal transcranial direct current stimulation reduces motor slowing in athletes and non-athletes. *BMC Neuroscience, 21*, 26.

Harris, D., Wilson, M., Buckingham, G., & Vine, S. (2019). No effect of transcranial direct current stimulation of frontal, motor, or visual cortex on performance of a self-paced visuomotor skill. *Psychology of Sport and Exercise, 43*, 368–373.

Kamali, A., Nami, M., Yahyavi, S., Saadi, Z., & Mohammadi, A. (2019). Transcranial direct current stimulation to assist experienced pistol shooters in gaining even-better performance scores. *The Cerebellum, 18*(1), 119–127.

Montenegro, R. A., Farinatti, P. T., de Lima, P. F., Okano, A. H., Meneses, A. L., de Oliveira-Neto, L., ... & Ritti-Dias, R. M. (2016). Motor cortex tDCS does not modulate perceived exertion within multiple-sets of resistance exercises. *Isokinetics and Exercise Science, 24*(1), 17–24.

Mizuguchi, N., Katayama, T., & Kanosue, K. (2018). The effect of cerebellar transcranial direct current stimulation on A throwing task depends on individual level of task performance. *Neuroscience, 371*, 119–125.

Parma, J., Profeta, V., Andrade, A., Lage, G., & Apolinario-Souza, T. (2020). TDCS of the primary motor cortex: Learning the absolute dimension of a complex motor task. *Journal of Motor Behavior, 53*(4), 431–444.

Rocha, K., Marinho, V., Magalhães, F., Carvalho, V., Fernandes, T., Ayres, M., Crespo, E., Velasques, B., Ribeiro, P., Cagy, M., Bastos, V., Gupta, D., & Teixeira, S. (2020). Unskilled shooters improve both accuracy and grouping shot having as reference skilled shooters cortical area: An EEG and tDCS study. *Physiology & Behavior, 224*, 113036.

Hikosaka, M., & Aramaki, Y. (2021). Effects of bilateral transcranial direct current stimulation on simultaneous bimanual handgrip strength. *Frontiers in Human Neuroscience, 15*, 674851.

**B. Unclear protocol and data**

Washabaugh, E. P., Santos, L., Claflin, E. S., & Krishnan, C. (2016). Low-level intermittent quadriceps activity during transcranial direct current stimulation facilitates knee extensor force-generating capacity. *Neuroscience, 329*, 93–97.

Lampropoulou, S., & Nowicky, A. (2013). The effect of transcranial direct current stimulation on perception of effort in an isolated isometric elbow flexion task. *Motor Control, 17*(4), 412–426.

Sasada, S., Endoh, T., Ishii, T., & Komiyama, T. (2017). Polarity-dependent improvement of maximal-effort sprint cycling performance by direct current stimulation of the central nervous system. *Neuroscience Letters, 657*, 97–101.

**C. No Sham condition /Article no available**

Andre, J., Vallence, A., Fujiyama, H., & Peiffer, J. (2019). Transcranial direct current stimulation does not enhance cycling time-trial performance.

**D. Cofounding factor**

de Sousa Fortes, L., Faro, H., de Lima-Junior, D., Albuquerque, M. R., & Ferreira, M. E. C. (2022). Non-invasive brain stimulation over the orbital prefrontal cortex maintains endurance performance in mentally fatigued swimmers. *Physiology & Behavior, 250*, 113783.

Nikooharf Salehi E, Jaydari Fard S, Jaberzadeh S, Zoghi M. (2022). Transcranial direct current stimulation reduces the negative impact of mental fatigue on swimming performance. *Journal of Motor Behavior, 54*(3), 327e36.
